# Self-paced silent speech brain-computer interface for device control

**DOI:** 10.1101/2025.04.09.25325542

**Authors:** Shiyu Luo, Miguel Angrick, Christopher Coogan, Daniel N. Candrea, Kimberley Wyse-Sookoo, Anouck Schippers, Rohit Ganji, Griffin W. Milsap, William S. Anderson, Chad R. Gordon, Donna C. Tippett, Nicholas J. Maragakis, Lora L. Clawson, Mariska J. Vansteensel, Francesco V. Tenore, Hynek Hermansky, Matthew S. Fifer, Nick F. Ramsey, Nathan E. Crone

## Abstract

Brain-computer interfaces can potentially restore autonomy to people with paralysis, including the ability to control their own environment via smart devices in the home. BCI applications for controlling smart devices to date have required visual displays or auditory cues, limiting the user’s ability to autonomously and privately issue commands. In this study, a clinical trial participant with amyotrophic lateral sclerosis (ALS) used a chronically implanted electrocorticographic (ECoG) BCI to control smart devices with self-paced silent speech commands. Across 18 experimental sessions, silently mimed speech commands were detected in real time and decoded with a median accuracy of 97.1% (chance: 7.14%). These results demonstrate that silently attempted speech can be reliably decoded without exogenous timing cues, supporting the feasibility of reliable autonomous smart device control with an implantable BCI.

## 1. Introduction

Neurological conditions such as amyotrophic lateral sclerosis can cause paralysis that impairs the abilities of affected individuals to communicate and to control their environment^1^. Existing alternative and augmentative communication (AAC) devices offer some remedy for restoring communication abilities, but their effectiveness is constrained for individuals who retain minimal or no residual muscle movement^2,3^.

Implantable brain-computer interfaces (BCIs) aim to assist these individuals by decoding attempted actions from the neural activities they generate in the brain^4–8^. Recent clinical trials of implanted BCIs have demonstrated the facilitation of a wide range of communication abilities, including textual communication and synthesized speech^6,9–14^. The accuracy and vocabulary of such systems have also steadily improved^15–21^. However, the most impressive of these demonstrations have relied heavily on the use of language models to correct decoding errors at the level of letters and words. In contrast to communication at the sentence level, efficient control of devices in the home, including computers, require reliable decoding of single-word commands with much higher accuracy than has been typical of sentence-level communication BCI’s. Thus, it remains to be determined whether speech BCIs can also be used to provide functional control over computers and other devices in the home, which is a critical unmet need for this population^22^.

Previous studies have shown that silently attempted speech can be decoded using implanted BCIs, but these approaches have required visual displays of timing cues in order to synchronize speech neural events and the decoding window^17,23^. This is not ideal for control-oriented BCIs where the display itself is not the primary media for feedback of decoding results nor the primary device that a user aims to control. The user also may want to turn on the display itself, creating a circular dependency on the very interface they are trying to control. Additionally, the requirement for a visual display from existing AAC and BCI technologies limits their use among individuals who have lost oculomotor control^24,25^.

Here, we report a clinical trial participant with ALS who used self-paced silent speech commands to control smart devices using a chronically implanted electrocorticographic (ECoG) BCI. Our participant’s BCI use, including initiation of the BCI, did not rely on a screen display. The decoder accurately detected the onset of self-paced silent speech and classified commands across 14 categories to control the operation of smart devices suitable for home use. Notably, this control was achieved without requiring decoder retraining or recalibration. This study provides a first proof of principle that an implantable speech BCI can be used to control smart devices directly and without requiring a screen display, potentially enhancing agency and autonomy of people living with severe motor impairments due to ALS and other neurological disorders.

## 2. Results

### 2.1 Overview of the neural assistant system

The study was conducted as part of the Corticom clinical trial (ClinicalTrials.gov Identifier: NCT03567213). A study participant with severe dysarthria and upper limb paralysis due to amyotrophic lateral sclerosis (ALS) was implanted with two 64-channel subdural ECoG grids over sensorimotor cortices (Figure 1a). Using neural signals from the lower grid covering areas activated during attempted speech, the participant was able to directly control smart devices by silently articulating commands such as “ light” , “ camera” , and “ television” at his own pace.

**Figure 1.**
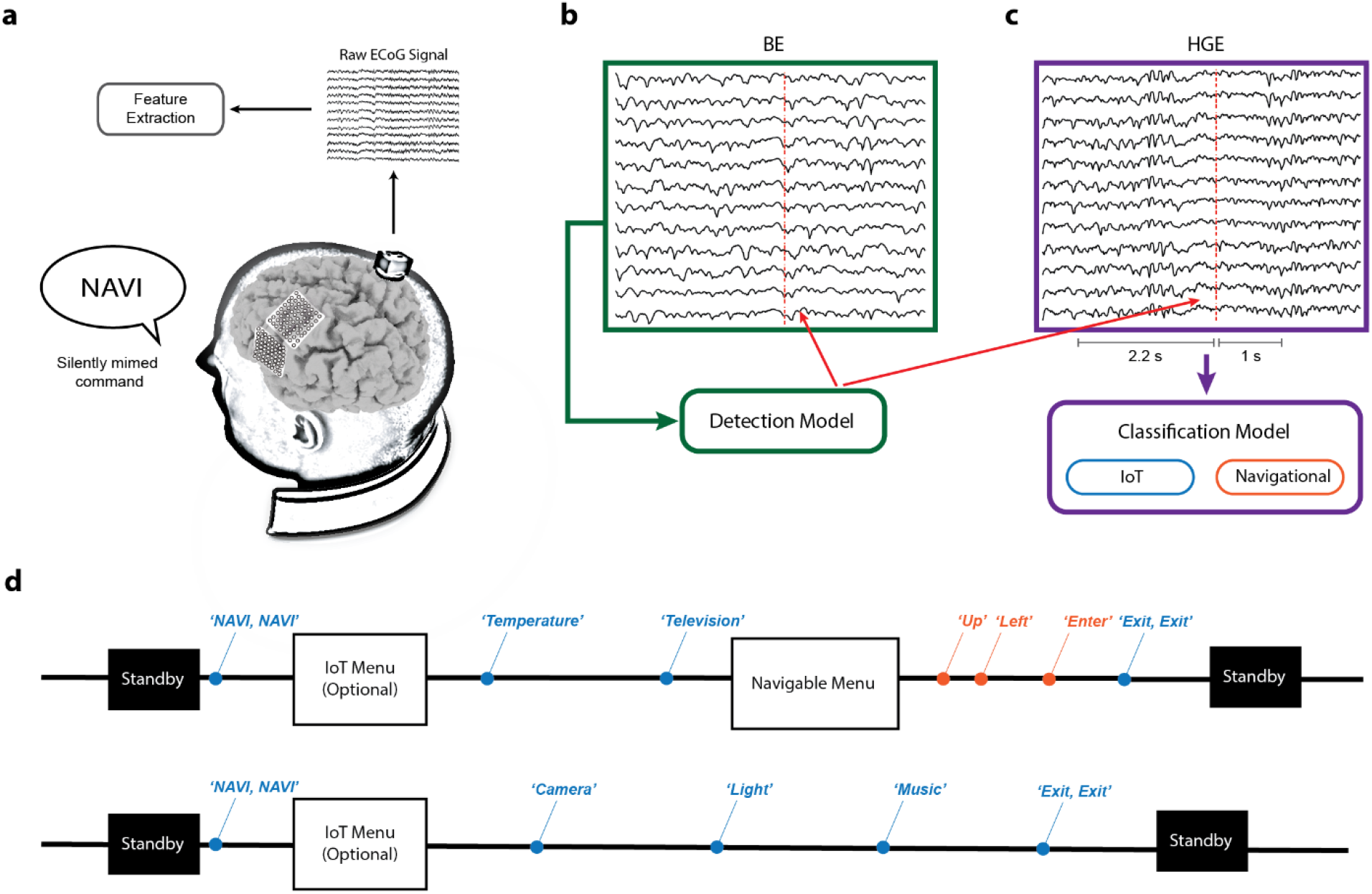
Schematic of the BCI system for smart device control. a) Neural signals during silently mimed speech were recorded by two 64-channel ECoG grids. b) Example of beta-band energy (BE, 13-30 Hz) for 12 channels. BE was used for detecting mimed speech events (dashed red lines) and segmenting the classification window. c) Example of high-gamma energy (HGE, 70-170 Hz) for 12 channels. HGE was used for training the classification model. The classification window was 2.2 s prior and 1 s after the event detection. d) Sample transcripts of the control commands by the participant.

While the participant was using the system, ECoG signals were streamed and processed in real time. Specifically, the signals were bandpass filtered between 13-30 Hz and 70-170 Hz to estimate beta-band energy (BE) and high gamma energy (HGE), respectively. Among ECoG BCI studies, the high gamma range is commonly used to generate neural features, because it is strongly correlated with neuronal spiking activities during movement^26–29^ and is highly stable^30^. The use of beta band for implantable BCIs has been less explored, but prior studies have shown that it is related to movement planning prior to execution as well as attention^31–33^. In this study, the timing of the participant’s speech intent was detected by identifying event-related desynchronization in the beta range (Figure 1b) while the content was classified using high gamma range features (Figure 1c). The neural assistant system was activated by silently repeating the keyword ‘NAVI’ two times. The system had two modes: an evaluation mode and a deployment mode. In the evaluation mode, a monitor displayed the decoded command. In the deployment mode, the system fulfilled the task associated with each command (e.g. turning on the light, the television, or opening a communication board application; Figure 1d). After activating the system by issuing two silent commands, the participant was able to control lights, stream music, alert caregivers, and get weather updates by issuing one of 14 silent Internet of Things (IoT) commands directly. Additional functions were supported by secondary menus that were navigable and controlled separately by using 6 silent navigational commands (Supplementary Note S1). For example, the ‘television’ command activated a smart TV application. TV channel selection was then accomplished by navigational commands (Supplementary Video S1).

### 2.2 Beta-band responses to detect silent speech

The speech intent of the participant was recognized by detecting event-related desynchronization in the beta-band. Not all channels exhibited robust event-related desynchronization corresponding to silent speech attempts (Supplementary Figure S2). We selected 15 channels that had consistent beta-band desynchronization for speech event detection (Figure 2a).

**Figure 2.**
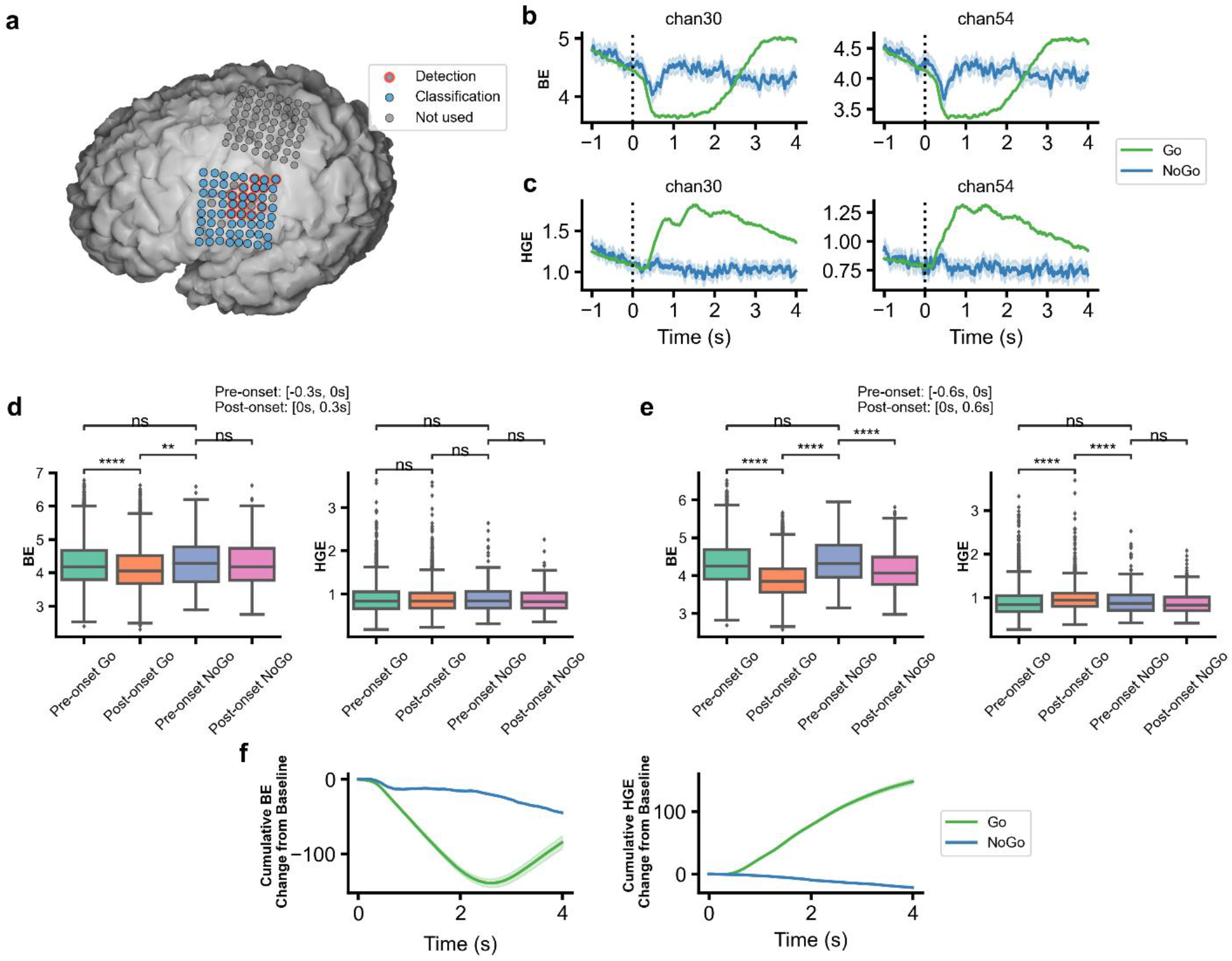
Comparison between BE and HGE for silent speech detection. a) ECoG grid location overlaid on top of MRI reconstruction of the participant’s brain anatomy. Blue electrodes are used in classification of the command identity. Electrodes with a red edge are used for detecting the silent speech attempt. Electrodes not used in this study are shown in grey. b) Average BE response in two example channels for all commands. Shaded areas represent 95% CI. *n*=3692 for Go trials, *n*=244 for NoGo trials. c) Same as b), but for HGE. d) Average BE and HGE 300 ms before and after stimulus onset under both Go and NoGo conditions. Each boxplot corresponds to *n*=41 experimental blocks (^****^p<0.0001, ^**^p<0.01, ^ns^not significant, two-sided Mann-Whitney U test with Bonferroni correction). e) Same as d), but for 600 ms before and after stimulus onset (^****^p<0.0001, two-sided Mann-Whitney U test with Bonferroni correction). f) Cumulative BE and HGE change from baseline averaged across trials. Shaded areas represent 95% CI. *n*=3692 for Go trials, *n*=244 for NoGo trials.

In a word reading task, we collected training data and evaluated the event-related activity changes in response to visual stimuli prompting the participant to silently produce the IoT words (Go condition) or to refrain from speech attempts (NoGo condition). In Go conditions, the participant saw the word he was supposed to silently pronounce on the screen; in NoGo conditions, he saw “ …” displayed instead and remained still. An evident decrease in BE was observed in both Go and NoGo conditions with a larger and more sustained decrease in the Go condition (Figure 2b). HGE saw a sustained and prominent increase in the Go condition while it remained mostly flat in the NoGo condition (Figure 2c). Intuitively, these results suggested that HGE might be sufficient for silent speech detection. However, we found that BE responded faster than HGE in the Go condition. In the Go condition, BE in the 300 ms window after stimulus onset was different from the 300 ms baseline prior to stimulus onset condition while HGE remained the same. Neither BE nor HGE responded to the NoGo condition during this time (Figure 2d). In the 600 ms after stimulus onset both BE and HGE changed from the 600 ms baseline prior to stimulus onset in the Go condition (Figure 2e).

Based on these results, we then investigated whether it was feasible to detect silent speech events using BE under the premise that these spectral responses changed during silent speech attempts and eventually returned to baseline. We found that the cumulative change from baseline (400 ms window prior to stimulus onset) in BE did decrease and subsequently increase under the Go condition. In contrast, in the 4 s after stimulus onset, cumulative HGE change kept increasing (Figure 2f).

### 2.3 Self-paced silent speech detection

Based on the results of our analysis of the training data, we ultimately used changes in BE to detect silent speech segments and changes in HGE for classification of silent speech commands in the resulting segments. Specifically, we identified the trough associated with the 1.8-s rolling average of BE in all detection channels. We then extracted HGE segments in all classification channels from 2.2 s prior to trough detection until 1 s after (Figure 3a). Note that detection accuracy was higher when the algorithm used BE (median: 96.11%, 95% confidence interval (CI): [93.33%, 96.67%]) than when it used HGE (median: 20.00%, 95% CI: [17.78%, 24.44%]) offline (p<0.0001, two-sided Mann-Whitney *U* test; Figure 3b).

**Figure 3.**
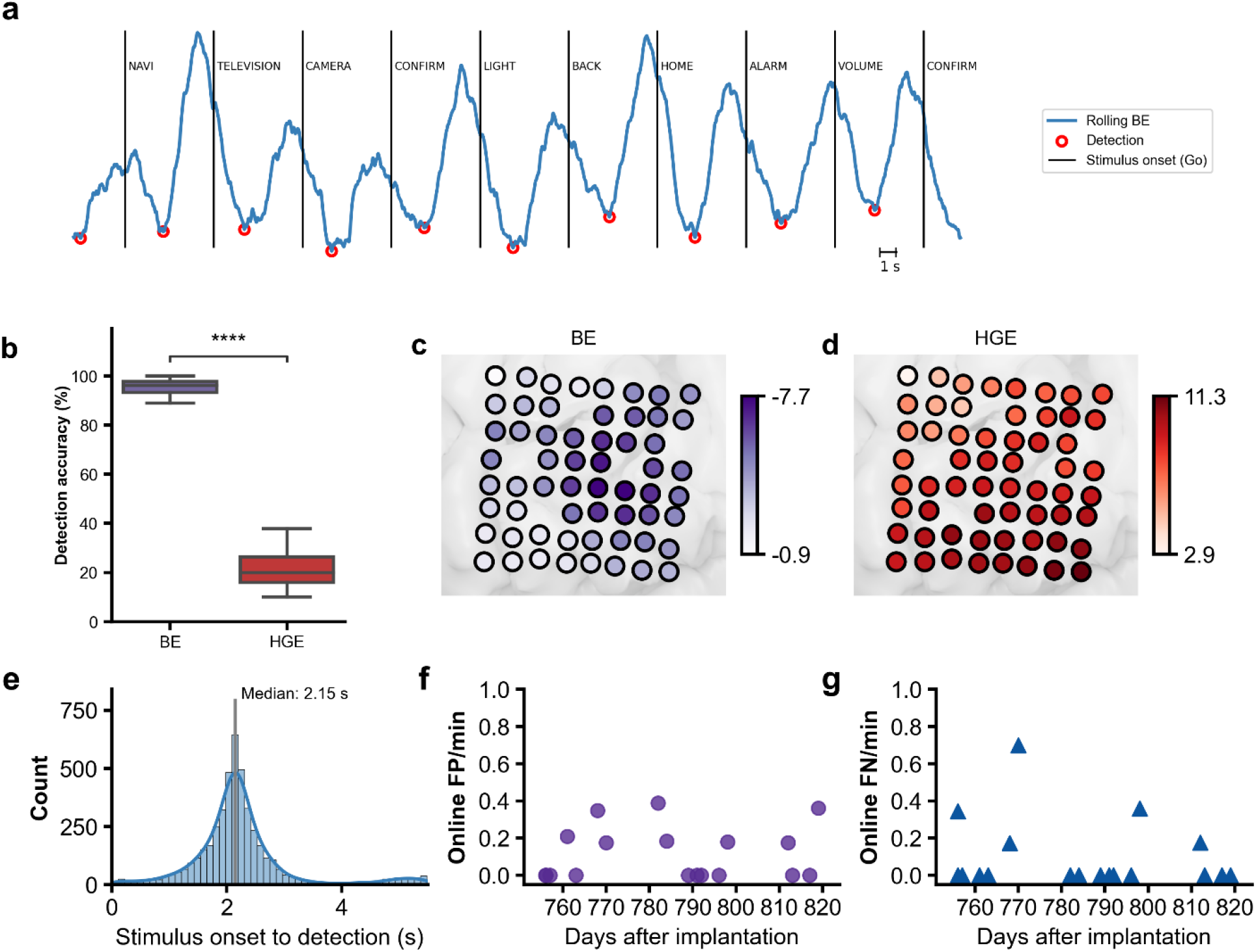
Silent speech detection performance. a) Illustration of the detection algorithm. Blue lines represent the 1.8-s rolling average of BE energy across all detection channels. Black lines indicate stimulus onset with the corresponding stimulus label shown to the right of the line. Red circles are the detected local minima b) Simulated detection accuracy during word reading tasks when the detection algorithm was applied to BE vs. HGE. Each boxplot represents *n*=41 experiment sessions (^****^p<0.001, two-sided Mann-Whitney U test with Bonferroni correction). c) Average BE at the time of detection for each classification channel, standardized against baseline (800ms to 0ms before stimulus onset). d) Same as c), but for HGE. e) Histogram of the interval between stimulus onset and detection using BE. f) Number of false positives (FP) per minute during closed-loop experiments. Each dot represents one real-time experimental session. g) Number of false negatives (FN) per minute during online usage. Each triangle represents one real-time experiment session.

The changes in band-limited energy had different cortical distributions for BE and HGE. We examined the z-scored BE and HGE at the time of detection in the training data (see Methods). More prominent BE changes were observed along the central sulcus in the dorsal part of the grid coverage (Figure 3c). HGE changes, on the other hand, were seen more notably in the ventral part of the grid coverage, close to the lateral sulcus (Figure 3d). We next investigated the pattern of the detection timing. The median interval between stimulus onset to detection was 2.15 s (95% CI: [2.14, 2.16]; Figure 3e). Given that our classification window was 2.2 prior to peak detection, we expected the majority of the segmented HGE to contain information starting from cue onset.

Finally, we examined the real-time performance of our approach in closed-loop experiments. In these experiments, the participant was instructed to silently mime one of the IoT commands at his own pace. The decoding results appeared on the screen as soon as they were detected. For detection, the median online false positive (FP) rate was 0 (95% CI: [0, 0.17]; Figure 3f) and the median online false negative (FN) rate was 0 (95% CI: [0.0, 0.18]; Figure 3g). These results indicated that BE spectral responses could indeed be used to support reliable self-paced silent speech decoding in real time.

### 2.4 Self-paced decoding performance

For direct device control, the classification accuracy of a BCI needs to be high. Since there is no intrinsic pattern to the commands being issued, this type of BCI cannot rely on language models to correct for decoding errors. In this study, we limited the vocabulary to 14 commands: *home, alarm, television, board, music, volume, light, message, exit, light, camera, temperature, NAVI, confirm*, and *back*. When evaluating the performance of the decoder, we instructed the participant to silently mime one of the 14 commands at his own pace. Decoding results were then displayed on the screen. The performance metrics were verified by the participant’s self-report and transcriptions from video. Specifically, if the decoding result did not match the silent commands, the participant attempted to make a grasp with his right hand to register the misclassification.

Online evaluation started 756 days, and ended 819 days, after implantation during 18 testing sessions. For the first day of online testing, two sessions were conducted. The remaining 16 days had one testing session each (Table S1). Across the study period, the median decoding accuracy was 97.10% (95% CI: [94.31%, 98.28%]; Chance: 7.14%, Figure 4a). The median correct decodes per minute was 11.86 (95% CI: [10.52, 12.60]; Figure 4c). Baseline was obtained during a single session conducted on Day 95 after device implantation and was used to calculate scores of HGE from both training data and testing data. No decoder retraining or baseline recalibration took place during the study period. Decoding performance remained stable with no statistically significant change observed (y = -0.03x + 116.08, where x is the days after implantation (same below), R^2^ = 0.02, p = 0.55; Figure 4a).

**Figure 4.**
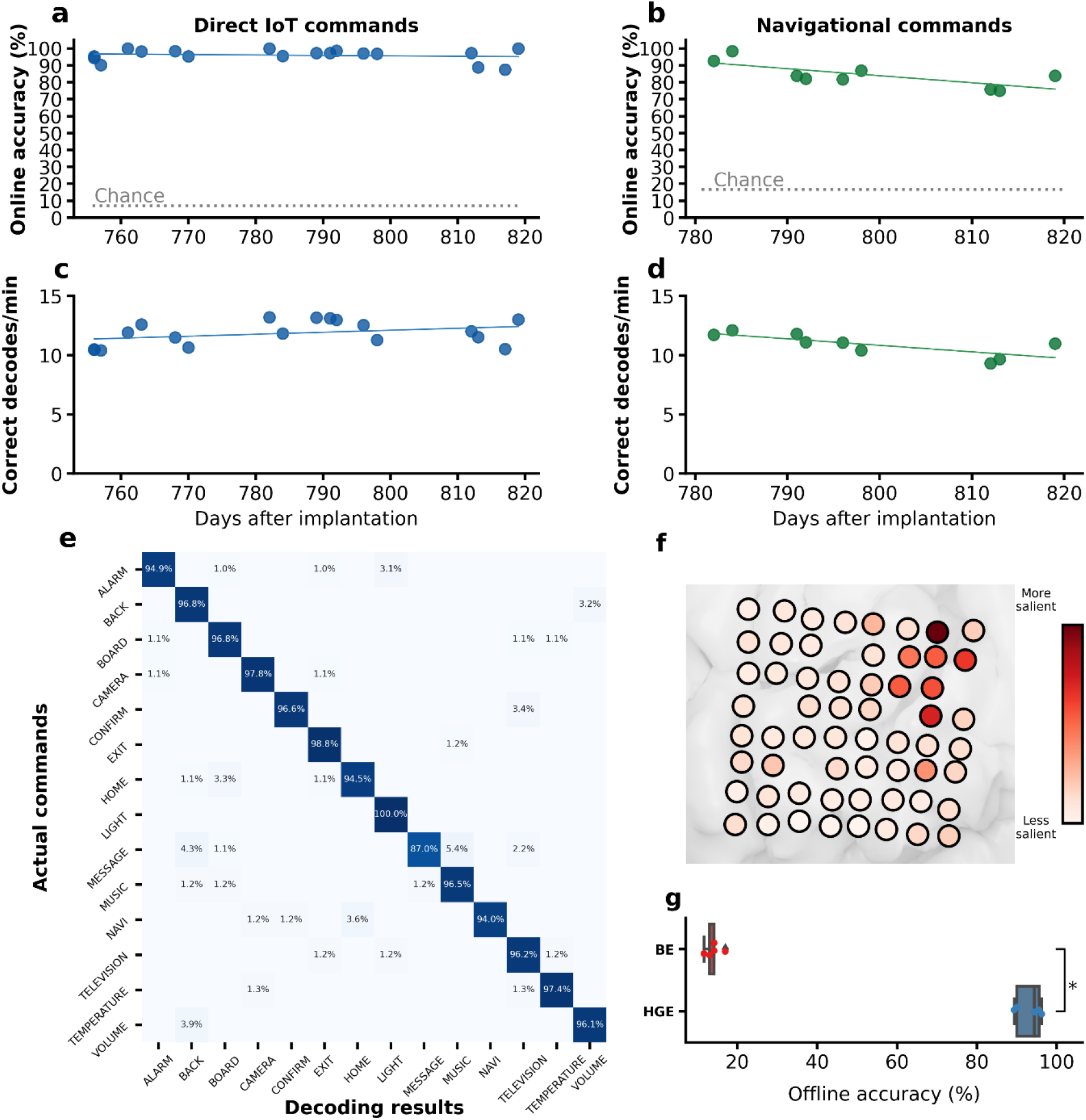
Self-paced decoding performance. a) Online accuracy of the direct IoT decoder. Each dot represents one experiment session. Chance = 7.14%. The solid line represents the linear least squares regression line between accuracy and days after implantation. b) Same as a), but for the navigational decoder. Chance = 16.67%. c) Number of correct decoding results per minute from the direct IoT decoder. Each dot represents one session. The solid line represents the regression line between correct decodes per minute and days after implantation. d) Same as c), but for the navigational decoder. e) Confusion matrix of all the direct IoT commands issued during online experiments. f) Relative electrode contribution to decoding performance for the direct IoT decoder. g) Accuracy of the IoT decoder after being trained with BE and HGE data offline (^*^p<0.1, two-sided Mann-Whitney U test with Bonferroni correction).

In addition to IoT control, the BCI system allows menu navigation via six navigational commands such as *up, down, left, right, enter*, and *back*. This navigational decoder shared the same architecture as the IoT decoder and was also controlled by the participant miming the command words silently. Online experiments using the standalone navigational decoder started 782 days and ended 819 days after implantation (Table S2). Across 9 online sessions, the median decoding accuracy was 83.78% (95% CI: [75.00%, 86.96%]; Chance: 16.67%, Figure 4b). No decoder retraining or baseline recalibration took place during the online evaluation period. Median correct decodes per minute were 11.06 (95% CI: [9.30, 11.71]; Figure 4d). A slight decrease in decoding performance was observed (y = -0.42x + 416.29, R^2^ = 0.54, p = 0.02; Figure 4b).

On Day 862 after device implantation, the participant attempted to control devices using both decoders (Supplementary Video S1). During this experiment, both the IoT decoder and the navigational decoder were active in the background, but only one decoder output was accepted by the system depending on the specific state of the application (Supplementary Note S1). For example, after the television was switched on, the system only accepted the output from the navigational decoder. The following actions were performed using the IoT decoder: shutting down the system, activating the system, turning on the music, turning off the music, turning on the light, and turning on the television. 8 out of 9 (88.89%) commands were correctly decoded.

While the participant was issuing the commands, the screen displayed a static menu of all 14 possible options. Once the television function was activated, the neural assistant system automatically switched to the navigational decoder. The participant selected from the visual interface of a commercially available smart TV application. The participant issued 13 commands to navigate across different menus and select one video to watch. 12 out of 13 (92.31%) navigational commands were correctly classified. On Day 866 after device implantation, the participant was instructed to control devices directly using the IoT decoder, this time without the options displayed on the screen (Supplementary Video S2). The IoT decoder correctly classified 12 out of 13 (92.31%) commands the participant issued to control the lights, trigger an alarm, get weather updates, confirm selections, and stream music.

The electrodes that had the greatest influence on decoding performance were located on the dorsal and posterior part of the grid compared to the ventral and anterior part of the grid for both direct IoT control commands and navigational commands (Figure 4f; Supplementary Figure S6). Although BE was found to be more useful for silent speech detection than HGE (Figure 2b), the classification accuracy based on BE is only slightly above chance (median: 14.15%) compared to that of chance accuracy (7.14%), and substantially lower than that based on HGE (median: 94.43%; Figure 4g).

## 3. Discussion

Among individuals living with ALS, home environment and computational device control are ranked as the most critical needs after communication assistance^22^. While able-bodied people can physically interact with their devices or utilize voice assistants that decode acoustic speech to interface with smart home devices, these options are often unavailable to those with severe paralysis. To address this gap, we developed a BCI that can allow direct device control for severely paralyzed individuals by decoding neural signals associated with silent speech commands. A clinical trial participant was able to activate a neural assistant and control devices such as a light and a smart TV by issuing silent commands at his own pace.

One outstanding question in speech BCI development is whether a visual display is necessary. If it is, then it may have limited utility for potential BCI users with impaired extraocular movements^24^. Additionally, some users could lose BCI control when the visual display is not available.

A key obstacle that has prevented BCI use without a visual display is that decoding algorithms have often relied on either visual or auditory timing cues to decode the neural signals associated with silent speech. Here, however, we show that beta-band spectral responses can be used reliably to detect silent speech attempts. However, we found that BE is not helpful for training classification models. Our findings are consistent with previous studies that have shown that beta-band power changes are sensitive to attention and motor planning, but do not contain information that can discriminate between spoken words or other movements of the same effectors^32,34^.

Historically, visual displays have played an important role in speech BCI development since they serve as the primary output for some communication-based BCIs. In ‘brain-to-text’ style BCIs^35– 37^, the output is usually displayed to both the user and their conversation partner^9,12^. However, for control-based BCIs, the output consists of device operations, which may not need to be presented on a screen. In this study, we introduced a new paradigm of speech BCI design that enabled direct control of smart devices without the need for screen displays. Another reason why a screen display is often an essential part of BCI applications is that training data collection typically takes place in experiments where visual stimuli are presented. For decoders that need decoder retraining or baseline recalibration before usage, this necessitates a screen setup prior to practical BCI usage. Here, in line with previous reports on the stability of ECoG signals, we were able to use a fixed decoder throughout the study while maintaining decoding performance. The fact that the same decoder could be used every day also allowed the participant to use the same decoder to activate the decoding system on each experimental day, supporting the feasibility of independent home use.

Although previous research has shown that covertly imagined speech can be decoded^38,39^, this study was entirely focused on decoding silently mimed speech. Further studies are needed to investigate whether covert speech (without articulatory movements) can be used to control devices directly. Another constraint of our study was the limited vocabulary. The IoT decoder had a vocabulary of 14 commands while the navigational decoder had 6 commands. The vocabulary size limited the control options available for the participant and made the control strategy of the neural assistant system less natural than that of a commercial voice assistant system. Whereas voice assistants are normally controlled by phrases such as “ Hey *assistant*, turn on the light” , our BCI system required the use of discrete in-vocabulary words such as “ NAVI, NAVI, light” .

Additionally, the results reported in this study were obtained from a single clinical-trial participant living with ALS. It remains to be determined whether the results will generalize to other people living with ALS or severe paralysis due to other neurological disorders. Another limitation of the study is the fact that the participant who took part in the study retained some level of articulatory movements during silent speech attempts in spite of significant weakness of articulators as a result of ALS. Further studies are needed to verify whether the same approach will still be applicable for people living with ALS who have lost all control over all articulatory muscles.

## Methods

### Clinical-trial overview

The study was conducted as part of the Investigation on the Cortical Communication (Corticom) System clinical trial (ClinicalTrials.gov; NCT03567213). Device use was approved by the US Food and Drug Administration under an Investigational Device Exemption. The participant gave informed consent for the study including the implantation of the device and all research associated activities. The Johns Hopkins Medicine Institutional Review Board (IRB0017247) approved the study protocol.

The pre-defined primary outcome of the clinical trial was to assess the safety and stability of the study device. These outcomes have been reported in previous publications^23,40^. In this study, the experimental paradigm and resulting data were not designed to directly report the primary outcome of the clinical trial.

### Participant

The study participant, a right-handed male, was in his 60s when the investigational device was surgically implanted and was diagnosed with amyotrophic lateral sclerosis (ALS) approximately 8 years before the start of the study. As a result of bulbar dysfunction, the participant had progressive dysarthria and dysphagia, accompanied by progressive dyspnea. All functional components of his speech were impaired, including respiration, phonation, resonance, and articulation.

### Neural implant

The neural implants for the participant were two 64-channel ECoG grids (PMT Corporation, Chanhassen, MN). They were surgically placed on the pial surface of the sensorimotor areas responsible for speech and upper limb movements in the left hemisphere. The ECoG grids were connected to an external digital head stage with a 128-channel percutaneous connector (Blackrock Microsystems, Salt Lake City, UT) anchored to the participant’s skull. Each ECoG grid had an 8×8 channel configuration with 4mm center-to-center spacing and a surface area of 12.11 cm^2^ (36.66 mm x 33.1 mm). Two reference wires were placed over anatomical regions not shown to be related to speech and upper limb movements and were used as reference for signal amplification. The electrodes were platinum-iridium discs with 0.76 mm thickness and 2 mm exposed surface diameter.

### Data collection

A NeuroPlex-E headstage (Blackrock Microsystems, Salt Lake City, UT), attached to the percutaneous connector, filtered (0.3-7500 Hz), amplified, and digitized the neural signals from the implanted ECoG grids. A mini-HDMI cable connected the headstage to a Digital NeuroPort Biopotential Signal Processing System (NSP, Blackrock Microsystems, Salt Lake City, UT). The NSP downsampled the signals to 2000 Hz.

### Real-time system

A PC connected to the NSP via a fiber optic cable recorded the neural signals. The signals were transmitted to a second PC from a ZeroMQ^41^ server implemented in BCI2000 signal processing module. The signal processing pipeline, the detection algorithm, and the classification model were implemented in Python with the ezmsg framework (https://github.com/iscoe/ezmsg) and deployed to this second PC.

### Peak detection in beta band

15 channels were selected from all the channels and used to support trough detection in the beta band associated with mimed speech. The channels were chosen based on their beta synchronization values and anatomical locations before online closed-loop experiments began. Neural signals acquired from these channels were bandpass filtered between 13 to 30 Hz. The logarithmic power of the filtered signals in 50 ms windows was computed every 10 ms. Detection features were computed from a rolling average of a 1.8-s buffer containing the beta logarithmic power. The troughs from a 3-s rolling buffer of the channel-averaged detection features were identified if the prominence was more than two times the standard deviation of the rolling buffer. To minimize false detections, there was a 2-s lockout period after each trough detection.

### Classification models

We trained convolutional neural networks (CNNs) on mimed speech from the word reading tasks. The detection method was applied to HGE from these tasks. We used the segmented periods from these neural features and the word labels to train the CNNs. Specifically, HGE segments in all classification channels from 2.2 s prior to trough detection until 1 s after were used for training. The model architecture we used was InceptionTime. Six Inception blocks were used, and each block had 3 Inception modules. In each module, there was an initial convolutional layer with 32 filters of kernel size = 1. Subsequently, 3 convolutional layers with kernel sizes ∈{5, 11, 23} and a MaxPooling layer with kernel size = 3 followed by a convolutional layer with 32 filters of size = 1 were used. The concatenated output from the 4 convolutional layers was the output from each module. The final output from the last Inception block was fed into a MaxPooling layer and, subsequently, a fully connected layer. The prediction from the fully connected layer provided the final classification score. An ensemble of 5 distinct neural networks were trained for both the IoT words and the directional keywords. The majority prediction from these models was adopted as decoding output. The model was implemented in Python 3.9 using PyTorch v 1.10.

### Experiment design and performance evaluation

When evaluating the performance of both the IoT and navigational decoders, the participant was asked to freely choose one of the words within the vocabulary. The application started with the instructions: “ Please choose an IoT word” , which remained visible throughout the duration of the experiment. The participant was instructed to mime the words without vocalization. The timing of the speech was determined by the participant. Once a speech event was detected and the decoding result became available, the decoded word appeared below the instruction on the screen and remained there until replaced by the next available decoding result.

The participant was instructed to make a grasp attempt with his right hand if the displayed decoding result was different than the mimed command. The attempted grasp movements were weak but visible. The ground-truth mimed command was transcribed by experienced personnel based on video recording of the participant’s face and hand movements.

During functional use, the participant controlled the whole system at his own pace. The system was activated with two consecutive silent *NAVI* commands. Afterward, a single command could trigger its corresponding functional control. The system could be turned off with two silent *Exit* commands.

### Statistical analysis

Performance data were presented as raw values. For statistical analysis, data outside 1.5 times the interquartile range were presented as outliers. Box plot whiskers were the maximum and minimum values of non-outliers. Statistical tests were performed in Python using Scipy packages.

## Data Availability

Performance data used to support the findings are available in paper or supplementary materials. Processed neural data will be made available upon manuscript acceptance. Raw neural recordings are available from the corresponding author upon reasonable request. They are not publicly available as they contain information that might compromise participant privacy.

## Author contributions

S.L. and N.E.C. conceived and designed the experiments. S.L. designed and implemented the detection and decoding algorithms. M.A., S.L., G.W.M. and C.C. implemented the real-time decoding pipeline. C.C. and S.L. designed and implemented the user interface. S.L., M.A., K. WS., and A.S. collected and analyzed the data. S.L., M.A., C.C., D.N.C., R.G., H.H., M.S.F., and N.E.C. contributed to the study methodology. D.C.T. performed speech and language assessment. N.J.M., L.L.C., M.J.V., F.V.T., N.F.R. M.S.F., and N.E.C. contributed to patient recruitment and regulatory approval. W.S.A., C.R.G., and N.E.C. planned and performed device implantation. H.H. supervised signal processing. N.F.R. and N.E.C. contributed to funding acquisition. N.E.C. supervised the study and the conceptualization. S.L. and N.E.C. prepared the manuscript. All authors reviewed and revised the manuscript.

## Acknowledgements

The authors would like to express their gratitude to participant CC01 and his family. The authors thank Alpa Uchil and the rest of the ALS clinic at the Johns Hopkins Hospital and the Johns Hopkins ALS Clinical Trials Unit for their care of the participant and their input on ALS-related topics; Mathijs Raemaekers and Kevin Nathan for fMRI data analysis; Yujing Wang for support in brain reconstruction and functional mapping; Ziwei Ouyang, Qinwan Rabbani, and Samyak Shah for help with data collection; Brock Wester for help with illustrations. The authors were supported by the National Institutes of Health under award number UHSNS114439 (NINDS).

## Notes

### Competing Interest Statement

The authors have declared no competing interest.

### Clinical Trial

NCT03567213

### Funding Statement

Research reported in this publication was supported by the National Institute Of Neurological Disorders And Stroke of the National Institutes of Health under Award Number UH3NS114439 (PI N.E.C., co-PI N.F.R.). The content is solely the responsibility of the authors and does not necessarily represent the official views of the National Institutes of Health.

### Author Declarations

The Institutional Review Board (IRB) of the Johns Hopkins Medicine gave ethical approval for this work and the Food and Drug Administration (FDA) gave approval under an investigational device exemption (IDE)

